# Factors associated with discrepancies in vaccination and immunization coverage of measles, tetani and hepatitis B among children aged 12-23 months in Narok County, Kenya

**DOI:** 10.1101/2025.03.04.25323382

**Authors:** Alfred Koskei, Fred Wamunyokoli, Mary Kerich, Simon Karanja

## Abstract

The burden of vaccine preventable diseases and mortalities are still unacceptably high in Narok County despite relatively high vaccination coverage. The objective is to determine factors associated with discrepancies in vaccination coverage and immunization coverage among 373 children aged 12-23 months. This was a cross sectional study that entailed determination of vaccination coverage and serosurveillance of Tetanus, Hepatitis B, and Measles using blood serum. A stratified sampling technique was used to sample children in the County. Statistical data analysis was done in SAS-software and regression performed using Pearson-chi.

**Results:** 378 children aged 12-23 months were sampled. The mean age of children was 15.3 (SD=3.7) months. Out of 378 children, 255 (67.5%) were aged <17 months. 195(51.6%) children were female. Children with normal birth weight (≥2.5kgs) were 260 (68.8%) while 230 (60.9%) received exclusive breastfeeding in the first six months of life. Up to 225(59.5%) children were fully vaccinated at health facilities while 31(8.2%) children were fully vaccinated at the outreach centers. Up to 127 (33.6%) children lived in houses with poor ventilations and hygiene. Children with acute/chronic infections during vaccination period were 34.1% (129/278) and 119 (31.5%) had helminthes infestation. Up to 249 (83%) children were vaccinated for measles dose one at the appropriate age of 9 month. Up to 119 (31.5%) children had previous hospital admission in the last one year. The Full vaccination coverage was 251(66.4%) while full immunization coverage was 38.1% (144/378). Discrepancies in vaccination coverage and immunization coverage per age and vaccine doses as were follows, respectively: MCV1 202(79.2%) versus 151 (59.2%) among children <17 months and 98 (79.7%) versus 78 (63.4%) among aged ≥17 months; Vaccination coverage for children who received both MCV1 & MCV2 was 68(55.3%) while immunization coverage was 50.4% among aged ≥17 months. The vaccination coverage for MCV2 was 8(6.5%) while immunization coverage was 3 (2.4%). For tetani vaccine: the vaccination and immunization coverage were 185(72.5%) and 133(52.2%) for those aged <17 months who received all 3 doses while those aged ≥17 months was 86 (69.9%) and 71 (57.7%, respectively. For children who received only two DTP vaccine doses, vaccination coverage was 32 (26%) and immunization coverage was 22(8.6%) among those aged <17 months while it was 6(6.5%) and 5 (8.6%) among aged ≥ 17 months, respectively. The main factors associated with discrepancies were birth weight ≥ 2.5kgs (OR=1.7), vaccination at health facility (OR=0.42), mixed breast feeding (OR=0.09), and no mother/ child history of helminthes infestation (OR=12.5).

## Introduction

Vaccination is a very effective health intervention in reducing child morbidity and mortality, averting 4.4 million deaths yearly (WHO/UNICEF, 2023). In various regions, many children remain unvaccinated, for example the vaccination coverage of diphtheria-tetanus-pertussis (DPT3) in 2023 was 84% globally, 95% in Europe and 69% in West-Central Africa, with estimated 25 million children globally not completing DTP doses in 2021 (WHO/UNICEF, 2023). In 2023, more than 14.3 million children under the age of one year did not receive any vaccine, also referred as zero dose children (WHO/UNICEF, 2023). Unfortunately, vaccines do not always provide immunity despite vaccinations. Globally, it is estimated that many vaccinated children are not immunized due to vaccine ineffectiveness and this include 77 million from tuberculosis following BCG vaccination, 19 million from measles, 18 million from poliomyelitis following vaccination with inactivated polio vaccine (IPV) and 10 million from pertussis and pneumococcus (Grassly, Kang & Kampmann, 2015). Some areas with high vaccination coverage have recorded outbreak of vaccine preventable diseases including measles among vaccinated children (Karami et al., 2017; Moghadam et al., 2014). According to Kenya National Measles Case Base Surveillance System, a total of 9,043 (Incident Rate=35/million) confirmed measles cases were recorded in Kenya between January 2003-December 2016, highest incident rate recorded in 2011 and 2012 and affected all counties in Kenya. The confirmed measles cases were recorded among 34% of infants who were vaccinated with at least one dose of measles virus containing vaccine (Ngina Kisangu et al., 2018). By 2016, Narok County was among the high burden counties in Kenya with measles incidence rate of 10-50/million that surpassed the measles elimination threshold target of 1 case/million despite vaccination coverage of 76% (Ngina et al., 2018). This could be partly contributed by discrepancies in vaccination coverage and immunization coverage.

### Methodology

This study was carried out in Narok County among 378 children aged 12-23 months between February 22^nd^ to May 26^th^, 2022. Narok County has 6 sub Counties namely Transmara West, Transmara East, Narok North, Narok East, Narok South and Narok West. It was a prospective cross-sectional study that involved the serosurveillance of measles, tetanus and hepatitis B vaccines among fully or incompletely vaccinated children. The assessment of uptake of measles, tetani and hepatitis B vaccines was done using semi-structured questionnaire administered to the child’s mother or guardian and their respective laboratory detection and quantification of vaccine-induced antibodies in blood serum done using indirect Elisa technique among fully or incompletely vaccinated children. The sample size (n) of 378 was determined using Fisher et al formula, 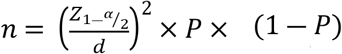 where the prevalence of fully vaccinated children was 58.5% with p=0.585 (KDHS. 2014) and the coefficient of the significance was 1.96 at level of 95% confidence interval; d=0.05 is the margin of error under the 95% confidence interval. The administrative wards in every sub County were sampled using proportionate stratified sampling. Then in every stratum (administrative ward), all households with child/children meeting the study inclusion criteria were physically identified and sampled using simple random sampling until desired sample size was attained in each stratum. Children with documentary evidence of immunization card or mother / child health booklet and have no known medical history or exposure to any vaccine preventable disease were eligible for the study. Children with history of blood and blood products transfusion and intravenous immunoglobulin’s receptor blockers in last three months were excluded in the study. Information on vaccination coverage was obtained from written immunization card or ANC health booklet. For every child sampled, his/her mother/guardian was asked to show the interviewer the vaccination card or ANC booklet where the child immunization data was documented. From the vaccination card or ANC booklet, the interviewer copied the dates of each vaccination received, the date of birth and age at vaccination, the birth weight and other health information therein. If the vaccination was not recorded in the vaccination card or ANC booklet and the mother/guardian is unable to show from any other document, it was considered not received even if corroborated by just oral explanation. The mother / guardian was also requested to provide any health booklet among those whose children had any medical history of disease or hospital admission to support their oral explanations related to child’s health as well as parental permission to give authorization for child’s blood withdrawal by the research assistant for serological analysis. The data to be copied include history of admission, previous medical history including specified disease/s the child was diagnosed with previously, history of helminthes infestation and place of vaccination (health facility and /or outreach) the child in question was vaccinated and place of delivery. The mother/guardian was to also requested to give oral details of the mode and duration of breast feeding for the sampled child in question, family history of disease and her/his experience with and perception of immunization services. The collection, processing and transportation of blood was carried out under standard good laboratory practices. Approximately 2.5mls of blood was collected through venipuncture by research assistants-all qualified phlebotomists from 25^th^ march 2022 to 17^th^ may, 2022. This involved preparation of site for injection using standard good clinical practices such as selection of site for vein puncture, cleaning the site by wiping with antiseptic alcohol and drying it and controlling pain and child trauma. The withdrawn 2.5ml blood in the syringe was transferred to appropriate laboratory equipment/specimen tube or a vacutainer with a red top with or without calcium citrate and stored in in a portable cold chain in -20°C. It was taken to the nearest health facility with centrifugation and refrigeration facilities to separate the serum using serum separation tubes with unique participant identifier and for cold storage. The blood serum was then stored at -20ºC, transported in dry ice by the principal investigator until it reached Metropolis pathology laboratory Eldoret for serological analysis. It was then stored at -70°C at the laboratory until it is analyzed. This was carried out under good laboratory practice and international regulations / guidelines. Indirect Elisa protocol was used to detect human IgG antibodies against the vaccine antigens (measles virus, tetanus and hepatitis B virus antigens). Ready to use specific IgG antibody level for measles, tetani and hepatitis B were studied using commercially available enzyme-linked immunoassay (elizas) kit sourced from Abcam plc (Cambridge, UK). The procedures were carried out as per the manufacturers’ instruction. Briefly, a 1.21 dilution of test samples were prepared by adding 10μl of the sample to 200 μl of the sample diluent. The negative and positive controls were supplied ready for use. 100μl of the controls and the blood serum samples was then dispensed to appropriate duplicate wells and incubated for 30 minutes at room temperature. The wells were washed three times with 1X washed buffer and blotted on an absorbent paper towel. This was followed by addition of 100 μl of enzyme conjugate and was then incubated for 20 minutes at room temperature. The wells were then washed three times with 300 μl of 1X wash buffer and blotted on absorbent paper towel. 100 μl of TMB substrate was then added to each well and incubated for 10 minutes at room temperature. This was followed by an addition of 100 μl of stop water followed by reading absorbance at 540nm against the reference filter of 600-650nm. Differences in optimal density (ΔA) were corrected by an internal control factor and used quantitatively to analyze the antibody presence.

A failure to seroconvert (negative or equivocal finding) following measles, tetanus or hepatitis B vaccination was considered as the inability to react to specific respective polysaccharide antigen. Samples with equivocal results were retested in duplicate. If similar results were obtained, samples were classified as equivocal. Otherwise they will be classified as positive or negative. The analyzed Eliza kit samples and the optical density measured with a photometer set at 540nm. The results of the Eliza were calculated using the kit instructions as adopted from the 3^rd^ WHO international standard guidelines. The quantitative titers were obtained from optical density values using the equation log10 titer=α*ΔA^β^ where α and β were specific constants for the kit lot. The cut off expressed in IU/ml were titers <9 IU/ml was considered negative, 9<x<11 was equivocal and ≥12 was positive for measles antibodies, while titers <0.1 IU/ml was considered negative, 0.1-<1.0 was considered equivocal/indeterminate and ≥1.0 IU/ml was considered positive (Immunized) for tetani antibodies. For Hepatitis B, Eliza test was performed and anti-hepatitis B surface antibodies (anti-HBs) binded to hepatitis B Surface Antigen (anti-HBsAg) coated on the test wells. The binding of peroxidase-labelled HBsAg conjugate to the anti-HBs completed the ‘sandwich’ formation and unwashed materials were washed away. Peroxidase catalyzes oxidation of luminogenic substrate, producing light which was detected and quantified. The intensity of the light is proportional to the amount of anti-HBs present in the sample. The anti-HBs <5.0 IU/ml was considered negative, ≥5.0≤x≤12 was equivocal / indeterminate and >12.0 was considered positive.

Statistical analysis was performed using SAS software version 9.4 (SAS Institute, Cary, NC, USA). Descriptive data on demographics variables and risk factors for vaccination and immunization coverage were presented in tables and or graphs in form of frequency counts, proportions, means and 95% confidence interval. The proportion of vaccination and immunization coverage was calculated for the general study population in relation to demographic, obstetric, vaccination status and risk factor groups. Inferential statistics on vaccination and immunization coverage was performed using Pearson’s Chi square tests on individual predictor variables and this informed univariate analysis. Fisher’s exact tests were performed for those variables with expected cell counts of less than 5. Threshold for significance level was set at <0.05%. Predictor variables with Pearson chi square statistic value of < 0.05 were subjected to univariate regression analysis. A few variables were dropped due to collinearity despite meeting this criterion. The final model was developed using backward elimination method. The study was approved by Moi University and Moi Teaching & Referral Hospital Institutional Ethics Committee (IREC) and the research permit was obtained from the National Council for Science, Technology and Innovation (NACOSTI) before commencement of the study. A formal letter of permission from Director of Medical services was obtained before the research process starts and was presented to all sampled medical superintendents and Facility-In Charges in Narok County. The parent/guardian’s permission were sought before any child was enrolled into the study.

## Results

The mean age of children was 15.3 (SD=3.7) months. Out of 378 children, 255 (67.5%) were aged <17 months while 123 (32.5%) were aged ≥17 months. 195(51.6%) children were female. Children with normal birth weight (≥2.5kgs) were 260 (68.8%). Out of 378 children, 230 (60.9%) received exclusive breastfeeding while 148 (39.1%) received mixed breast feeding in the first six months of life. Out of 378 children, 347 (91.8%) children were vaccinated at a health facility and 31 (8.2%) children were vaccinated at various outreach centers. Out of 251 fully vaccinated children, 249 (99.2%) were vaccinated at various health facilities while 2 (0.8%) fully vaccinated children were vaccinated at outreach centers. Out of 300 children vaccinated specifically with measles vaccines dose one, 44 (14.7%) children were vaccinated at level 2 health facility, 144 (48.0%) children at level 3 health facility, 100 (33.3%) children at level 4 health facility, 12 (4.0%) children at level 5 health facility while 31 (6.9%) children were vaccinated at outreach centers. Up to 129 (34.1%) children had documented medical history of acute or chronic infections during or shortly before vaccinations period and 119 (31.5%) were admitted at various hospitals in varied period within the first year of their childhood life. Children with documented medical history of helminthes infestations during vaccinations period were 119 (31.5%).

Two hundred and fifty-one (66.4%) children lived in homes with acceptable good ventilation and hygiene standards while 127 (33.6%) children lived in houses with poor ventilations and hygiene standards. Out of 378 children, 306 (81.0%) were delivered at a health facility while 72 (19.0%) children were delivered at home with no skilled birth attendant. Up to 347 (91.8%) children were vaccinated at health facility while 31(8.2%) were vaccinated at an outreach center. These and other immunization characteristics were summarized in context of their immunity status in table 1.

**Table 1.**
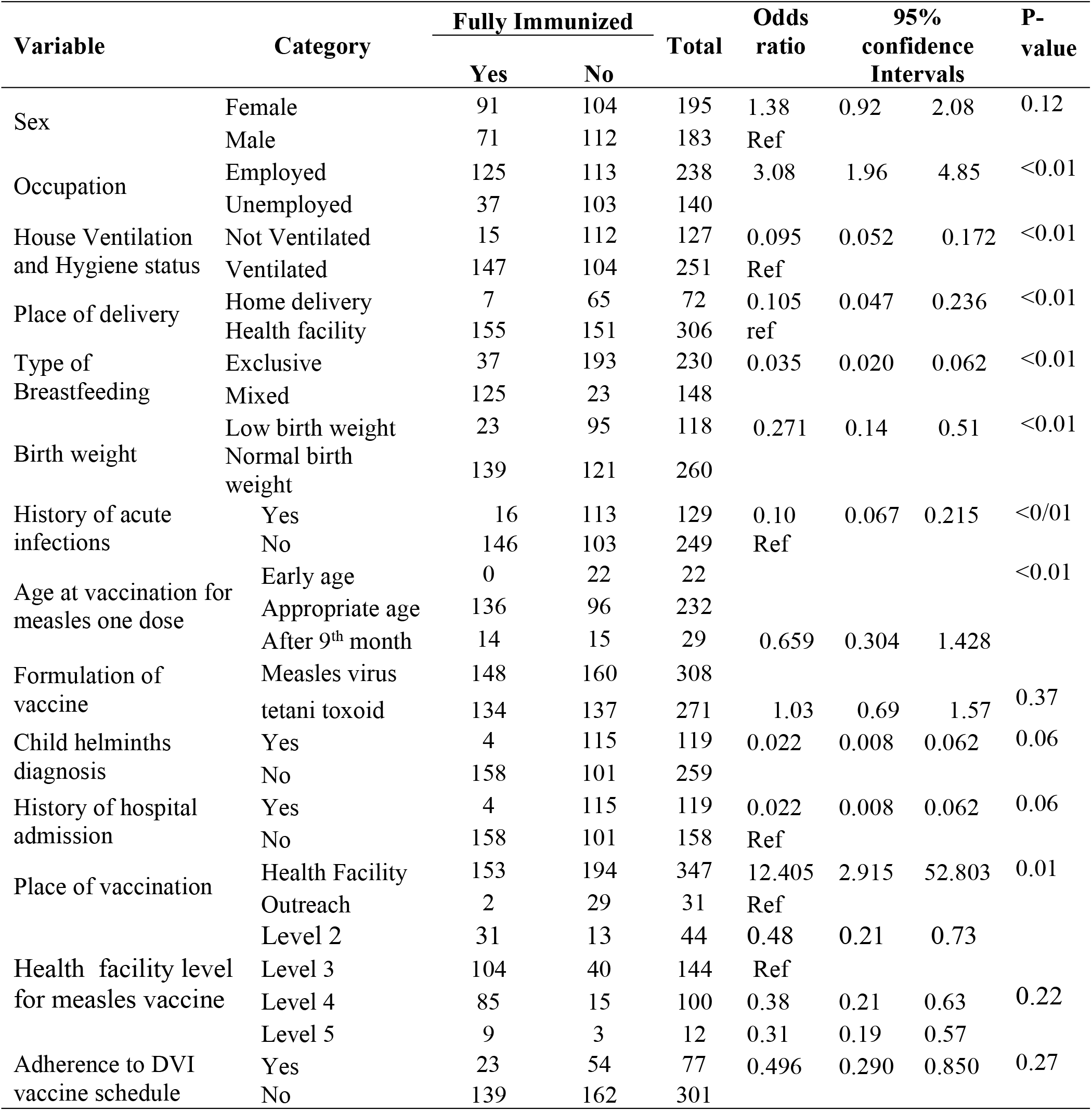
Immunization characteristics per immunity status.

Out of 378 children, 339 (89.7%) children received pentavalent dose one, 351 (92.9%) children received pentavalent dose two and 324 (85.7%) children received pentavalent dose three, 300 (79.4%) children received measles virus containing vaccine dose one and out of 123 eligible children, 76 (61.8%) received measles virus containing vaccine dose two. 251 (66.4%) children were fully vaccinated for basic antigens and this included 172 (67.5%) fully vaccinated children aged <17 months (inclusive of measles one dose only) and 79 (64.2%) fully vaccinated children aged ≥ 17 months (inclusive of measles one and two doses).

The study revealed discrepancies between vaccination coverage and immunization coverage that may indicate the gaps in effectiveness of childhood immunization program. While the fully vaccinated children who received all the age specific DVI scheduled vaccines were 251 (66.4%), the proportion of fully immunized children were 144 (38.1%). The age appropriate vaccination coverage for measles dose one was 202 (79.4%) and for all doses one and two was 68(55.3%) while the overall immunization coverage for measles was 235 (62.2%); (<17 months: 154 (65.5%) and ≥17 months: 81(34.5%). Among children aged <17 months, while vaccination coverage for measles dose one was 202 (79.2%), the immunization coverage was 151(59.2%). A consistent trend in other singular measles doses was noted among children aged ≥ 17 months. For children who received two doses of measles one and two, the vaccination coverage was 68 (55.3%) while the immunization coverage was 62 (50.4%). Similarly, for tetani the proportion of vaccinated children that received all the three essential doses of pentavalent vaccines was 271 (71.7%); (<17 months:185(72.5%) and ≥ 17 months: 86 (69.9%) while the proportion of immunized children was 204 (54%) (<17 months: 133 (52.2%) and ≥17 months: 71 (96.8%); (<17 months: 248(97.3%) and ≥17 months: 118 (95.9%) while the proportion of immunized children was 262 (69.3%) (<17 months: 169 (66.3%) and ≥17 months: 93 (75.6%).

The proportion of children vaccinated against hepatitis B that received all the three essential doses of pentavalent vaccine was 271 (71.7%); (<17 months:185 (72.5%) and ≥ 17 months: 86 (69.9%) while the proportion of children immunized against hepatitis B were 219 (57.9%); (<17 months: 146 (57.3%) and ≥17 months: 73 (28.6%). Similarly, the proportion of children vaccinated against hepatitis B that received at least two essential doses of pentavalent vaccine was 366 (96.8%); (<17 months:248 (97.3%) and ≥17 months:118 (95.9%) while the proportion of children immunized against hepatitis B were 290 (76.7%); (<17 months: 195 (76.5%) and ≥17 months: 95 (77.2%). The discrepancies in vaccination and immunization coverage were summarized in table 2.

**Table 2.**
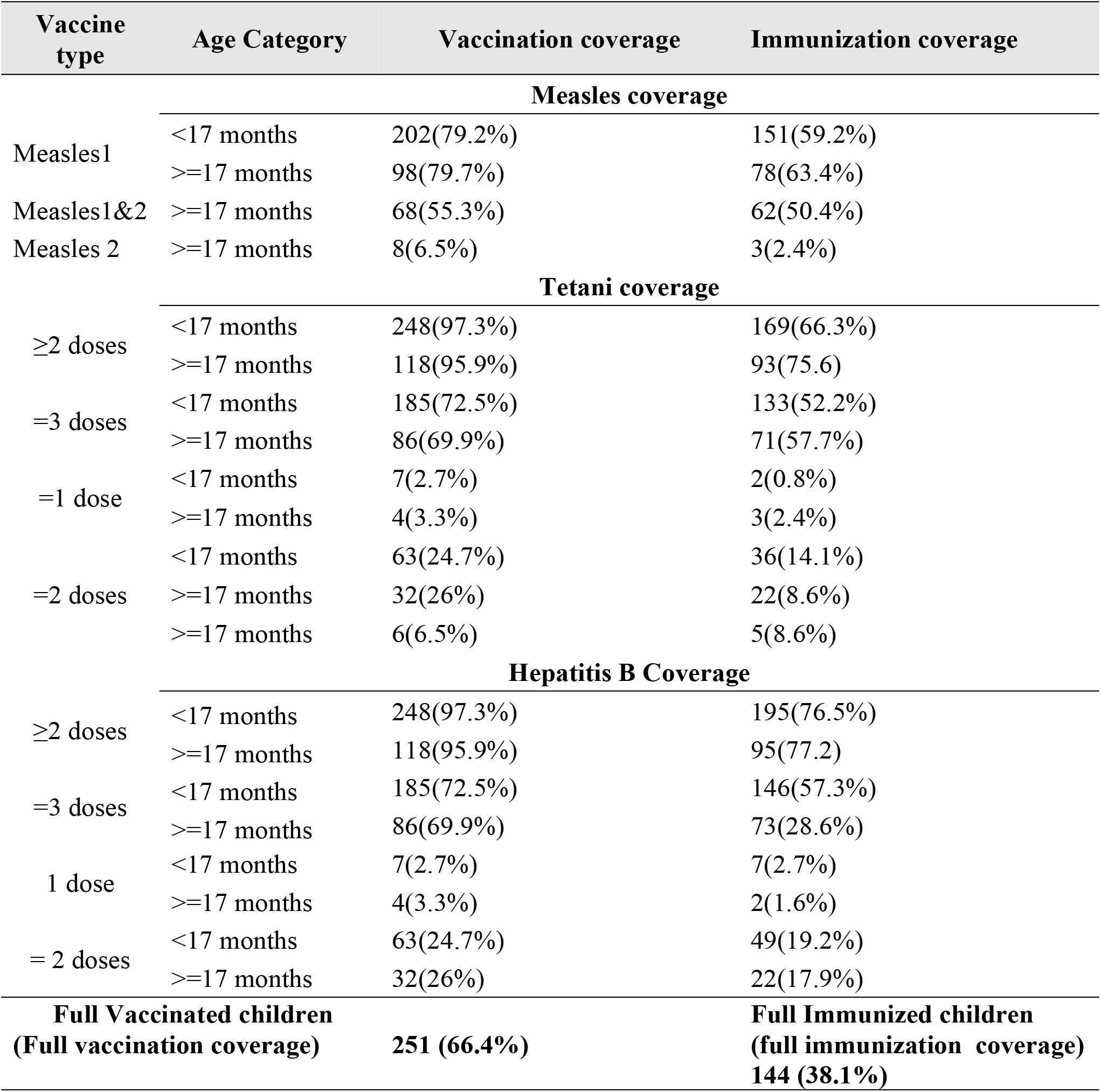
Discrepancies between vaccination and immunization coverage per age.

Multivariable regression analysis was performed for predictor factors that showed a significant association (using cutoff point of p<0.05) with outcome variable at univariate analysis. Sex, house type, age at vaccination and adherence to immunization schedule were not associated (p-value>0.05) with discrepancies in vaccination and immunization coverage hence were dropped from the model. Controlling for other variables in the model, variables such as occupation (p-value=0.38), child history of acute infection diagnosis (p-value=0.27), birth weight (p-value=0.08), ventilation / hygiene status (p-value=0.62), and place of vaccination (p-value=0.29) were not statistically significant with discrepancies in vaccination and immunization coverage. Place of delivery, mother/child helminthes infestation and type of breastfeeding were significantly associated with overall immunization. Home delivery significantly reduces the odds of being immunized when vaccinated compared to health facility deliveries. Children who were delivered at home were 74% (OR = 0.26) less likely to be immunized compared to children who were delivered in a health facility (p-value=0.029). Exclusive breastfeeding significantly reduces the odds of being immunized compared to mixed breastfeeding. Children who experienced exclusive breast feeding were 91% (OR = 0.09) less likely to be immunized compared to children who experienced mixed breast feeding (p < 0.01). Children who had no history of helminthes infestation were 12.5 times (OR =12.5) more likely to be immunized compared to children with history of helminthes infestation (p = 0.003). These were summarized in table 3.

**Table 3.** Multivariate Analysis.

| Table 6. Multivariate Analysis |  |  |  |  |  |  |
| --- | --- | --- | --- | --- | --- | --- |
| Immunized overall | Odds ratio | std. err. | z | P>z | [95% CI] |  |
| **Birth weight** |  |  |  |  |  |  |
| Normal birth weight (Ref) |  |  |  |  |  |  |
| Low birth weight | 1.7198 | 1.316107 | 0.71 | 0.479 | 0.383828 | 7.706616 |
| **Occupation** |  |  |  |  |  |  |
| Employed | 0.38 | 0.19 | -1.89 | 0.058 | 0.142 | 1.034 |
| Unemployed (ref) |  |  |  |  |  |  |
| **House type** |  |  |  |  |  |  |
| Mud Wall (ref) |  |  |  |  |  |  |
| Stone Wall | 2.23 | 1.53 | 1.17 | 0.24 | 0.58 | 8.55 |
| Timber Wall | 2.77 | 1.59 | 1.78 | 0.08 | 0.91 | 8.51 |
| **Place of vaccination** |  |  |  |  |  |  |
| Health Facility | 0.42 | 0.35 | -1.04 | 0.29 | 0.08 | 2.16 |
| Outreach (ref) |  |  |  |  |  |  |
| **Place of delivery** |  |  |  |  |  |  |
| Home delivery | 0.26 | 0.16 | -2.18 | 0.03 | 0.08 | 0.87 |
| Health Facility (ref) |  |  |  |  |  |  |
| **breastfeeding type** |  |  |  |  |  |  |
| Exclusive | 0.09 | 0.04 | -6.30 | 0.01 | 0.05 | 0.21 |
| Mixed (ref) |  |  |  |  |  |  |
| **Child's history of acute infections** |  |  |  |  |  |  |
| No | 1.76 | 0.89 | 1.12 | 0.27 | 0.65 | 4.72 |
| Yes (ref) |  |  |  |  |  |  |
| **Age at vaccination** |  |  |  |  |  |  |
| Appropriate age (ref) |  |  |  |  |  |  |
| Late vaccination | 0.67 | 0.42 | -0.63 | 0.53 | 0.19 | 2.27 |
| **Mother-child helminth diagnosis** |  |  |  |  |  |  |
| No | 12.52 | 10.62 | 2.98 | 0.003 | 2.376 | 66.007 |
| Yes (ref) |  |  |  |  |  |  |
| **Ventilation/hygiene status** |  |  |  |  |  |  |
| Not Ventilated | 0.6180 | 0.4196 | -0.7100 | 0.4780 | 0.1633 | 2.3384 |
| Ventilated (ref) |  |  |  |  |  |  |
| **have problem with adherence to immunization schedule** |  |  |  |  |  |  |
| No | 1.2981 | 0.6721 | 0.5000 | 0.6140 | 0.4705 | 3.5810 |
| Yes (ref) |  |  |  |  |  |  |
| cons | 0.6240 | 0.8088 | -0.3600 | 0.7160 | 0.0492 | 7.9156 |
| **Level of health facility** |  |  |  |  |  |  |
| Outreach (Ref) |  |  |  |  |  |  |
| Level 3 | 0.174478 | 0.229609 | -1.33 | 0.185 | 0.013231 | 2.300929 |
| Level 4 | 0.211729 | 0.287418 | -1.14 | 0.253 | 0.014801 | 3.028792 |
| Level 5 | 0.356433 | 0.5384073 | -0.68 | 0.495 | 0.018459 | 6.882636 |
| cons | 76.06965 | 147.25 | 2.24 | 0.025 | 1.712059 | 3379.902 |

## Discussion

The results indicated that 38.1% of children were immunized while 66.4% were fully vaccinated. This is far below the recommended 94% herd immunity required to contain measles outbreak, given basic reproduction number for measles (R_0_=16). These findings were also below the WHO’s set minimum standard threshold of >95% for attainment of universal immunization coverage among children in Narok County, Kenya (WHO, 2021). This indicate a high unmet need for effective vaccination services and high vulnerability of children to vaccine preventable diseases. The low immunization coverage (38.1%) in context of the vaccination coverage (66.4%) indicate the ineffectiveness of measles, tetani and hepatitis B vaccinations.

Discrepancies in vaccination and immunization coverage have been documented by other studies (Izadi et al., 2015; Polonsky et al., 2015; Hayford et al., 2013; Cutts, Izurietta & Rhoda, 2013; Nigatu et al., 2008). The extent of this gap is influenced by many factors ranging from child (host) factors (age, age at vaccination, health status, nutritional status) to cold chain programming (Ataguba et al., 2016; Knoll et al., 2014; Cherry, 2012; Enquselassie et al., 2003; Knoll et al., 2014; Bruguera et al., 1990). These findings are consistent with the present study findings that found discrepancies in the proportion of fully vaccinated children (66.4%) and fully immunized children (38.1%).

The main factors associated with discrepancies in vaccination and immunization coverage were: mixed breastfeeding, helminthes infestation and place of delivery. The present study demonstrated that children who experienced exclusive breast feeding were 91% (OR = 0.09) less likely to be immunized after vaccination compared to children who experienced mixed breast feeding (p < 0.01). This could be attributed to interference by high titer levels of residual maternal antibodies among children who were exclusively breastfed for at least six months (Metcalf et al., 2016; Ion-Nedlcu et al., 2001). The present study findings are therefore consistent with many studies that have documented that exclusive breast feeding decrease vaccine effectiveness and thus associated with discrepancies in vaccination and immunization coverage especially in measles vaccine (Zahrael et al., 2016; Enquselassie et al., 2003; Akande et al., 2007; (Leuridan & Van Damme, 2007; Gans et al., 2001; Klinge et al., 2001; Redds et al., 2004; Metintas et al., 1997; Yeager, Davis & Ross, 1977; Albrecht et al., 1977). However, it contradicts Wei Wang et al study in China that found no association between breast feeding status and vaccine effectiveness (Wei Wang et al., 2024). It also contradicts Borras et al., 2011 that found no significant association between maternal antibodies and measles immunization coverage among children aged 9 to 14 months in Catalonia (Borras et al., 2011).

Birth weight of a baby was not statistically significant factor (p-value=0.479) affecting immunization coverage. Low birth weight is partly a reflection of malnutrition and this finding is consistent with Wei Wang *et al*., and Zahrael et al. studies that found no relationship between immunization coverage and malnutrition (Wei Wang *et al*., 2024; Zahrael et al., 2016). This is similar to Sultana et al., study finding that showed that the immunization coverage was 42% among children with good nutritional status and 26.5% among children with poor nutritional status, with no significant statistical differences (Sultana et al., 2006). However, it contradicts many studies that have documented that children with low birth weight attributed to malnutrition including underweight had higher disparities in vaccination and immunization coverage than those with good nutrition status (Prendergast, 2015; Kizito et al., 2013; Patel et al., 2009; Akande et al., 2007; Kielmann et al., 1976; el-Gamal et al., 1996; Marcos et al., 1997; Brussow et al., 1995; Dao et al., 1992).

There was no association between the mother’s education level and full immunization coverage (p value>0.05). This study postulate that mothers’ education level lack plausible biological mechanism towards immunologic response of children to vaccination against measles, tetani’s or hepatitis b vaccines. Social relationship could exist as parents’ education is an important risk factor for delaying the vaccination and this could make sense in context of measles vaccine. This contradict Zahrael et al., that found that there is a relationship between mothers’ education level and children’s’ immunological responses and that children of mothers who had below 9 years of formal education were five times more likely to be immunized for measles compared to those children whose mothers had at least 9 years of formal education (Zahrael et al., 2016). It is also similar to Sultana et al. Bangladeshi study that found no association between parental education level and immunization coverage (Sultana et al., 2006).

The present study assessed the economic and environmental conditions using the type of houses and their respective ventilation and crowding status that children routinely expose themselves in their homes before, during and after vaccination. The study revealed that children from higher economic background with better environmental conditions such as those living in stone wall houses had no significant difference in immunity status compared to children from low economic / poor environmental conditions’ households living in mud or unfurnished timber wall houses (p-value=0.24). Similarly, children who live in poorly ventilated households had no statistical differences in immunity status compared to children who lives in well ventilated households with good hygiene (p=0.48). This could reflect on their parent’s employment or occupational status (p-value=0.06). These contradicted many studies that have documented that poor environmental health conditions were associated with high disparities in vaccination and immunization coverage as well as higher vaccine failure rate (Naylor et al., 2015; Patel et al., 2009). Another study had demonstrated changes in transmission patterns of measles after an outbreak that was significantly associated with poor environmental health conditions related to overcrowding in Harare, Zimbabwe (Marufu et al., 2001). The present study contradicts many other studies and this could be due to differences in the study designs and eligibility criteria used as the present study had sampled healthy children with no prior exposure to any vaccine preventable diseases or viral antigen or any history of infectious disease outbreak.

The present study found that child’s age at vaccination for measles was not a significant factor affecting disparities in immunization coverage (p-value=0.53). However, the present study showed an increase in proportion of disparities in coverage among children (n=3; 15%) vaccinated early for measles virus containing vaccine dose one than those vaccinated on appropriate time (n=195; 78.3%) or late vaccination after age 9^th^ month (n=19; 65.5%). The present study finding is consistent with the American and Korean academy of pediatrics guidelines that do not recommend early vaccination unless there is/are indication/s for early vaccination/s such as presence of community outbreak involving infants with ongoing risk for exposure or before departure for international travels to an area with endemic and endemic levels of disease (American academy of Peadiatrics, 2015.pg 536-47; Korean Paediatric Society, 2015. Pg 141-62). It also supports Black *et al* recommendation for contraindication of early administration of measles vaccines on grounds that it reduce vaccine efficacy among children with poor developed immune system (Black et al. 1984) as well as Hasley et al., study recommendation that measles vaccination should be delayed, done after eight months of age among children in Haiti after he carried a study that showed that seroconversion rate increased from 45% to 100% from 6^th^ month to 12 months (Hasley et al., 1985). However, the present study finding contradicts studies that have documented age at vaccination as a determinant to immunization coverage (Carazo et al., 2020; Nic Lochlainn et al., 2019; Ataguba et al., 2016; Knoll et al., 2014; Defay et al., 2013; Poethko-Muller & Mankertz, 2011). It also contradicts a cohort study by Kanida et al., conducted in Niger that showed that immunization coverage increased with age at vaccination, from 78% in single dose given at the sixth month to 95% at the nine month (Carazo et al., 2020; Kanida et al., 1998). Furthermore, present study contradicts Hammarlund and de Melker’ study on tetanus antibody durability that showed that age at vaccination influenced the durability of tetanus antibody titers in serum (Hammarlund et al., 2016; de Melker et al., 2000). The present study found that place of delivery was associated with disparities in coverage (p-value=0.03) and that home delivery significantly reduces the odds of being immunized (OR=0.26) by 74% compared to hospital facility’ deliveries after vaccination. This could be attributed to intervention by professional skilled attendant in inoculation of vaccines (esp. tetani and hepatitis b) and utilization of complementary health services in an integrated postnatal care system with child’s immunization and nutritional supplementation which descreases discrepancies. It could also be attributed to indirect effect of cold chain programming such as health facilities’ readiness to vaccinate that has direct impact on vaccine utilization unlike a non-facility exposure/home delivery (LaFond et al., 2014; Naimoli et al., 2008; Haddad et al, 2009) as well as hygienic practices associated with deliveries by skilled personnel such as doctor, nurse or/ and midwife especially in tetani toxoid vaccine (Kyu et al., 2013; Birmingham et al., 2004).

The present study found no significant association between the mode of delivery and discrepancies (p-value >0.05). This was contradicted by Wei Wang study that found that in measles virus containing vaccine, the infant born via caesarean section was 2.2 times less likely to be immunized compared to infants born by vaginal delivery (Wei Wang et al., 2024). Some studies have postulated that caesarean section deliveries have long term effects on immune response particularly for pneumococcal, meningococcal conjugate vaccines and hepatitis B inactivated vaccine thorough early exposure to life gut microbiota’ which could have triggered early infant immune maturation (de Koff *et al*,. 2022; Lin et al., 2020).

Absence of mother or child’s helminthes infestation significantly increase the odds of being immunized / reduce discrepancies compared to those with history of helminthes infestation. The present study showed that children who had no history of helminthes infestation were 12.5 times (OR =12.5) more likely to be immunized compared to children with history of helminthes infestation (p = 0.003). This is attributed to pathophysiologic changes associated with parasitic infestation that weakens the nutritional and immunologic health of the child’s host. This finding is consistent with other studies that have documented that helminthes predisposes children to milder infections that reduce vaccine effectiveness, hence affecting high discrepancies (Akande et al., 2007; Marufu et al., 2001; Lyamuya et al., 1999; Kambarami et al., 1991). However, it contradicts the findings of two studies that found no effect of helminthes infestations and discrepancies in immunization coverage (Kampmann & Jones, 2015; Elliot et al., 2010).

Child’s medical history of acute or chronic infections was not statistically significant (p-value=0.27) with discrepancies in coverage. This finding is consistent with studies that showed that childhood infections do not affect antibody responses and immunization coverage on children (Kampmann and Jones, 2015; Jones et al., 2011; Elliot et al., 2010; Akande, 2007). It is also similar to many studies that showed no difference in antibody responses to MMR vaccination between children with diarrhea (Halsey et al., 1985; Ndikuyeze et al., 1988), fever (Halsey et al., 1985; Ndikuyeze et al., 1988), afebrile upper respiratory tract infections (Cilla et al., 1996) and healthy children. The present study contradicts some studies that have documented discrepancies in vaccination and immunization coverage among children with the following infections and conditions: high parasitemia levels and HIV in pregnancy (Hatgers, 2008; Cumberland et al.,2007), upper respiratory infections (Usen et al., 2000), chronic diarrhea after OPV vaccination (Parker et al., 2014; Haque et al., 2014), non-polio enteroviruses at the time of vaccination (Parker et al, 2014), HIV after hepatitis B vaccination (Gouvea et al., 2015), HIV after BCG vaccination (Elliot et al., 2010; Lule et al., 2015), HIV after measles (Kizito et al., 2013; Miles et al., 2008) and tetani vaccinations (Eliott et al., 2010) and acute leukemia after vaccination for diphtheria, tetanus and inactivated polio vaccine (Lawrence et al., 1976) as well as during post-organ transplant (Balloni et al., 1999). However, clinical trials and epidemiological studies have showed that malaria chemoprophylaxis or HIV antiretroviral agents do not reduces tetani or measles vaccine antibody response in vaccinated children and are both safe for use in children with such conditions, irrespective of the immunocompetence status (Rosen & Breman, 2004; WHO, 2018; Roper et al., 2017). The present study finding also contradicts other studies that found that constant exposure to chronic infections or infections that occur concurrent to measles vaccination adversely affected antibody responses in children (Naylor et al., 2015; Patel et al., 2009; Kambarami et al., 1991; Aaby et al., 1986; Migasena *et al*., 1998; Krober et al., 1991). The present study found that children whose parents or guardians had poor adherence to immunization schedule had no statistically significant difference in immunization coverage discrepancies (p-value=0.61) compared to those with good adherence. Although there is no direct biological plausible explanation for this, indirectly it may affect the child’s age at vaccination including delays in vaccination which has been shown to boost measles antibody responses but seems has no effect on tetani and hepatitis b vaccine responses (Hasley et al., 1985) but has also been contradicted by many other studies (Ataguba et al., 2016; Knoll et al., 2014; Isik et al., 2003). Vaccine dosages intervals, which is dependent on programming of immunization schedule, may impact negatively on immunization coverage if inappropriate dosage interval is experienced by the child owing to failure to adhere to the routine childhood immunization programming schedule (Wei Wang et al., 2020; Cherry, 2012; Knoll et al., 2014). Also, vaccine spacing of doses in infant vaccination schedule that have longer intervals between doses have been demonstrated to have higher vaccine induced antibody responses than vaccines in short-spaced vaccination schedule (Halsey & Galazka, 1985; Booyet et al., 1992). This could be related to pharmacokinetic and pharmacodynamics interactions that could reduce vaccine effectiveness.

Place of vaccination (p-value=0.29) was not a significant factor affecting discrepancies in immunization coverage despite data showing 58% less likelihood probability of being immunized (OR=0.42) if vaccinated at an outreach center compared to a health facility. The present data also showed the trend that immunization coverage was higher among children vaccinated at health facilities (78%) compared to those vaccinated at outreach centers (6.5%) specifically for measles vaccines. The present study postulates this insignificance to the pastoralism and migratory nature of the study population that enabled ‘cross over’ of children to different health facilities in the County, based on their location at the time of seeking vaccination services. It was common to observe in the vaccination health records that majority of vaccinated children obtained different vaccines doses in various health facilities. Also, some children who got vaccines at outreach centers also obtained other vaccines at other health facilities of different levels and due to lack of standardization of place of vaccination in this study, it neutralized the effect of this factor in influencing immunization coverage. It was easy to expect that immunization coverage would be lower in outreach centers compared to higher level facilities due to potency risk of vaccines associated with cold chain’s temperature changes, vaccine exposure to light and tampering as well as other environmental risks that could occur at the site or during transportation. These findings therefore contradict studies that documented that place of vaccination and transportation influences the vaccine viability in general and immunization coverage in particular (Kamanda et al., 2010; Akande, 2007; Oyefolu et al., 2007; Samant et al., 2007; Weir, 2004).

The significance of the formulation of vaccine as a factor influencing immunization coverage was not demonstrated in this study (p-value=0.37) as there was no significant difference in the proportion of immunization coverage in single type, live attenuated measles RNA vaccine (n=148; 48.1%) and tetani toxoid vaccines (n=134; 49.4%) among children in Narok County. This contradict some studies that found that Vaccine that contains single type of vaccine such as measles tend to illicit better intrinsic immunological profiles than multiple types vaccine formulations (Center for disease control, 2014; Milstein &Gibson., 1990). This finding indirectly confirms the hypothesis that measles virus is phenotypically mono typic hence they have one serotype / epitope which can protect against exposure to any or all the 24 viral genotypes (Tahara et al., 2013) while tetani toxoid is highly antigenic (Lodha et al., 2000) hence partly explain similarities in immunization coverage in different doses. It also justifies the present study methodology as tetani infection lacks sub-clinical phase of the disease hence no other non-vaccine induced antibodies (including IgM) could erroneously be detected and quantified that was induced by wild tetani clostridium or infection other than the vaccine. Both vaccines are co-administered with other respective antigens and its effects on immunization coverage could not be distinguished in this study. Hence it does not affirm Philiph *et al*., findings that showed that vaccine formulations influences seroconversion and immunization coverage (Philips et al., 2017). It also contradicts Boyet et al., study that showed that vaccine such as hepatitis B that have been formulated to contain a high optimal concentration may have better priming and higher antibody responses if given in high doses (Boyet et al., 1992).

## Conclusion

The study found high discrepancies (>10%) between vaccination coverage and immunization coverage in the three tested basic antigens (measles, tetani and hepatitis B) among children in Narok County, Kenya. The full immunization coverage was 38.1% (n=144/378) while full vaccination coverage was 66.4% (n=251/378).

History of child helminthes infestation (p-value=0.003), exclusive breast feeding (p-value=0.01) and home birth deliveries (p-value=0.03) were associated with discrepancies in vaccination coverage and immunization coverage. Child’s history of acute infections, place of vaccination, age at vaccination, economic status, birth weight and formulation of vaccines were not associated with disparities in vaccination and immunization coverage.

## Data Availability

The original data set in excel of this study is available upon request.

## Ethical Clearance

The study was approved by Moi University and Moi Teaching & Referral Hospital Institutional Ethics Committee (IREC) and the research permit was obtained from the Kenya National Council for Science, Technology and Innovation (NACOSTI) before commencement of the study. The parent/guardian’s permission were sought before any child was enrolled into the study.

## Declaration of Conflict of Interest

I declare I don’t have any financial, social or any other such conflict of interest

## Acknowledgement

I thank Dr Erick Mibei of University of Kabianga for reviewing this manuscript especially statistical and serological analysis. I grateful to Dr. Mathew Koech of Oak Tree Healthcare and Professor Wamzandi of University of Kabianga for financial support. I thank all laboratory personnel of Metropolis Pathology Laboratory in Eldoret.

## Notes

### Competing Interest Statement

I declare that i didn't have any competing conflict of interest

### Funding Statement

The financial cost of this study was supported by the research grant mostly from the University of Kabianga Directorate of Research and author's personal savings.

### Author Declarations

The study was approved by Moi University and Moi Teaching & Referral Hospital Institutional Ethics Committee (IREC) and the research permit was obtained from the National Council for Science, Technology and Innovation (NACOSTI) before commencement of the study. The Joint Board Ethics Approval Committee of Moi Teaching and Referral Hospital and Moi University is a leading institutional Ethics Review Board in Kenya, established in 1998 under the delegated mandate from The Kenya National Commission for Science, Technology and Innovation (NACOSTI). The Board have twelve (12) executive committee members drawn from various institutional departments and local community comprising of physicians and other medical practitioners, scientists, biostatisticians, lawyers, sociologist and anthropologist, religion expert and community representative/layperson. The chair is periodically rotating between the two institution and has a board secretary and backed up human subject administrator and a number of support staff. Every month, the board approves up to 75 protocol applications on human subject research ranging from clinical trials to epidemiological studies, among others, which are applied through an e-portal. All members are trained on human subject protection and are distinguished bioethicists on their own standing. It is funded by the two institutions and also charges a fee on every proposal/protocol application for approval.

